# Genome-wide analyses of variance in blood cell phenotypes provide new insights into complex trait biology and prediction

**DOI:** 10.1101/2024.04.15.24305830

**Authors:** Ruidong Xiang, Yang Liu, Chief Ben-Eghan, Scott Ritchie, Samuel A. Lambert, Yu Xu, Fumihiko Takeuchi, Michael Inouye

## Abstract

Blood cell phenotypes are routinely tested in healthcare to inform clinical decisions. Genetic variants influencing mean blood cell phenotypes have been used to understand disease aetiology and improve prediction; however, additional information may be captured by genetic effects on observed variance. Here, we mapped variance quantitative trait loci (vQTL), i.e. genetic loci associated with trait variance, for 29 blood cell phenotypes from the UK Biobank (N∼408,111). We discovered 176 independent blood cell vQTLs, of which 147 were not found by additive QTL mapping. vQTLs displayed on average 1.8-fold stronger negative selection than additive QTL, highlighting that selection acts to reduce extreme blood cell phenotypes. Variance polygenic scores (vPGSs) were constructed to stratify individuals in the INTERVAL cohort (N∼40,466), where genetically less variable individuals (low vPGS) had increased conventional PGS accuracy (by ∼19%) than genetically more variable individuals. Genetic prediction of blood cell traits improved by ∼10% on average combining PGS with vPGS. Using Mendelian randomisation and vPGS association analyses, we found that alcohol consumption significantly increased blood cell trait variances highlighting the utility of blood cell vQTLs and vPGSs to provide novel insight into phenotype aetiology as well as improve prediction.

## Introduction

The complete blood count is amongst the most routinely ordered clinical laboratory tests performed globally^1^. Blood cells play crucial roles in a variety of biological processes, such as oxygen transport, iron homeostasis, and pathogen clearance^2–4^, and serve as key biological conduits for interactions between an individual and their environment. The genetic architecture of blood cell traits has been recently elucidated by genome-wide association studies (GWAS)^5,6^ and, consistent with their well-known role in disease and clinical testing, blood cell traits are both highly heritable and have been genetically linked to many diseases, including cardiovascular diseases^7^, mental disorders^8^ and autoimmune diseases^9^.

Despite the success of GWAS, our understanding of the genetic architecture of complex traits has been limited by a focus on mean trait values and how these change with respect to genotype. The genetics of trait variance, how individual measurements deviate from the mean trait value across genotypes, is far less studied. It has long been known that trait variance, e.g. for gene expression^10,11^ and metabolic rate^12^, plays a role in an organism’s fitness and phenotypic penetrance. Theories support the existence of selection on trait variance to improve fitness ^13,14^. However, there are limited observations of selection on clinically significant traits. Variance quantitative trait loci (vQTLs) have been identified for human body composition traits, such as BMI^15,16^, and for cardiometabolic biomarkers^17^. vQTLs have also been linked to gene-by-environment interactions (GxE) or gene-by-gene interactions (GxG)^15–18^. vQTL studies of blood cell traits are currently lacking, despite their central role in biological processes and ubiquity in clinical testing.

Polygenic scores (PGS) are being intensively studied in various ways to determine their utility in clinical practice^19–21^. PGS for blood cell traits, in particular, are both highly predictive and show sex- and age-specific interactions^6,7^. How to treat trait variance and vQTLs with respect to phenotype prediction is relatively unexplored. A variance PGS (vPGS) to predict the trait variance may be estimated from the effect sizes obtained from a genome-wide vQTL analysis. In theory, a PGS is different from a vPGS, where the former may be used to stratify individuals based on the inherited trait level while the latter stratifies individuals based on the inherited deviation of individuals from the population mean. It is known that the accuracy of a PGS varies across individuals as a function of the genetic distance from the reference population^22^. As a vPGS may represent the outcome of GxE^16^ or GxG due to the nature of vQTLs^15^, examining a PGS alongside vPGS may reveal individual variability in PGS accuracy that can be accommodated.

Here, we conduct genome-wide vQTL analysis for 29 blood cell traits in the UK Biobank^6,7^ and the INTERVAL cohort^23^. We compared the discovered vQTL with conventional QTL and analysed vPGS with conventional PGS in the prediction of blood cell traits. We found novel vQTL, not identified by previous conventional GWAS and displayed strong selection to reduce blood cell trait variances. Finally, we demonstrate the use of vPGS in stratifying individuals, resulting in differing PGS performance, and then show that PGS performance within vPGS strata is associated with lifestyle factors.

## Results

### Genome-wide discovery and annotation of vQTLs in the UK Biobank

We performed GWAS of variance in 29 blood cell traits from the UKB^17,18^ (Average sample size = 402,142, **Supplementary Table 1**). The processing of phenotypes and genotypes followed previously established protocols with stringent quality control and normalisation procedures^5–7^. Levene’s test^24^, as implemented in OSCA^25^, was used to map vQTLs for each of the 29 blood cell traits and the inflation factors and lambda GC were assessed using LD Score regression (LDSC)^26^. Across the 29 traits, the average lambda GC and LDSC intercepts were 1.03 and 1.007, respectively (**Supplementary Table 2**), indicating negligible inflation. At a study-wide significance level of p < 4.6×10^-9^ and with clumping *r*^2^ < 0.01, we identified 176 independent vQTLs (**Figure 1a**, **Supplementary Table 3**, **Methods**).

**Figure 1.**
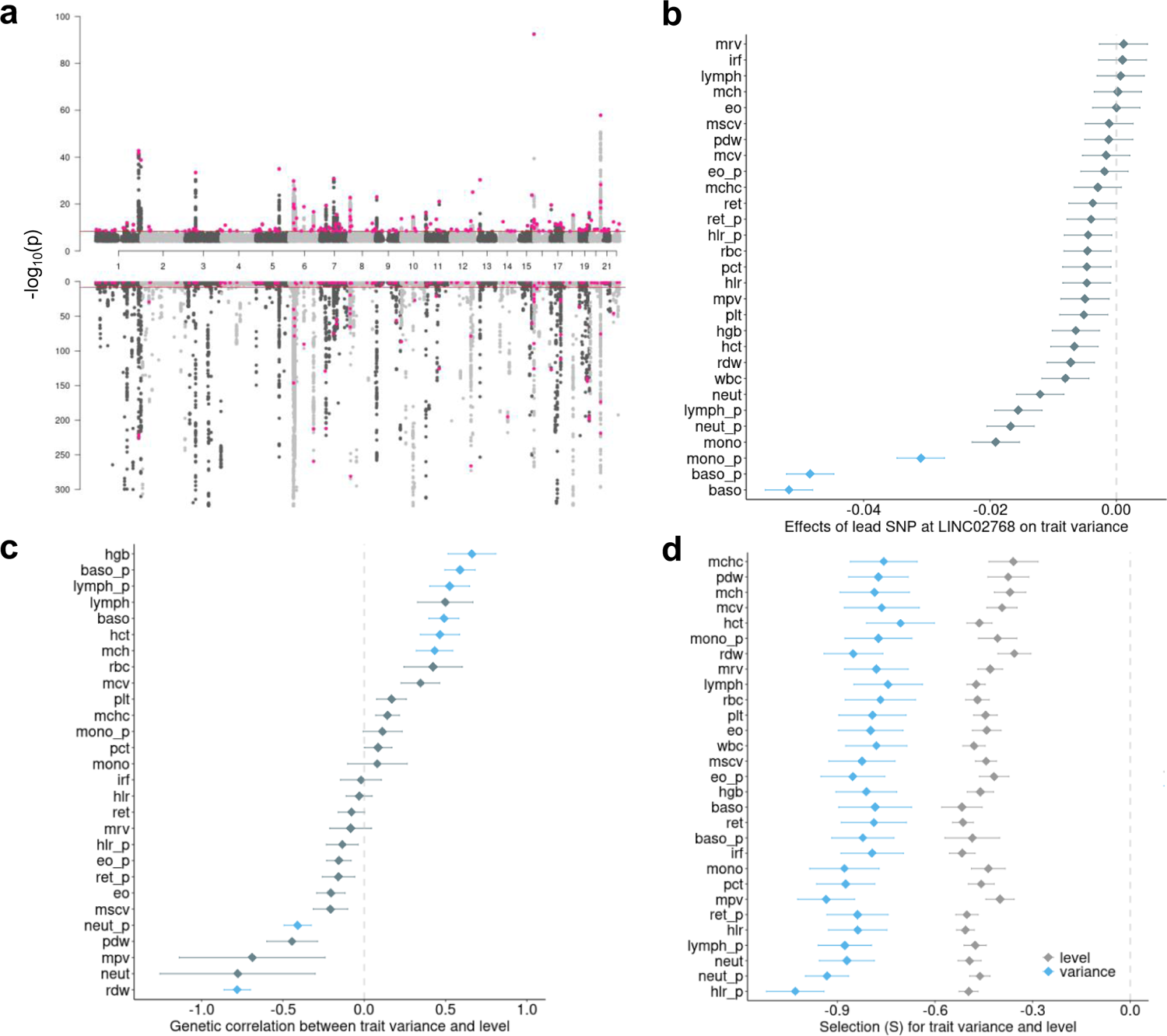
vQTLs for 29 blood cell traits and their comparison with additive QTLs. **a**: Miami plot showing the best (smallest p-value) vQTLs across 29 blood cell traits (top plot) and the corresponding best additive QTLs (bottom plot). Red dots are genome-wide significant independent vQTLs. **b**: Example of pleiotropic effects of the C allele of rs10803164 for the long non-coding RNA *LINC02768* on blood cell trait variance. Blue indicates the effect on trait variance had p < 4.6×10^-9^ (study-wide GWAS significance). **c**: Genetic correlation between blood cell trait variance and trait level. Blue indicates the correlation had multi-testing adjusted p < 0.05. **d**: Selection coefficient estimated by BayesS ^28^ for trait variance and level.

Basophil cell count (baso) and basophil percentage of white cells (baso_p) yielded the largest number of independent vQTLs (N = 27 and 23, respectively), whereas high light scatter reticulocyte count (hlr) did not have any study-wide significant vQTLs (**Supplementary Table 4**). Most vQTL were associated with the variance of only one or two traits and many of these traits are correlated (**Supplementary** Figure 1 **and Supplementary Table 3**). By counting the number of blood cell traits associated, the most pleiotropic lead vQTL was located in gene *HBM* (hemoglobin subunit mu) and was associated with the variance of four traits (red blood cell count, mean corpuscular volume, mean corpuscular hemoglobin and mean corpuscular hemoglobin concentration, **Supplementary Table 3**). The second pleiotropic lead vQTL related to long intergenic non-coding RNA *LINC02768* was associated with 3 traits [monocyte percentage of white cells (mono_p), baso and baso_p, **Figure 1b**]. To account for the phenotypic correlations, the pleiotropy of trait variance was further assessed using HOPS^27^, which found that 495 SNPs (out of 71,216 input SNPs) showed significant pleiotropy (**Supplementary Table 5**). In this analysis, the most significant pleiotropic locus was *LINC02768* (**Supplementary Table 5**). vQTLs were largely distinct from additive QTLs. Of 176 lead vQTLs, 147 were not detected as additive QTLs by Vuckovic et al^6^, the largest GWAS to date of blood cell traits. vQTLs had an average r^2^ of 0.33 (SD=0.12) with the lead additive QTLs from Vuckovic et al^6^ (**Supplementary** Figure 2). We repeated the OSCA^25^ analysis fitting the trait level as a covariate, i.e. effects of vQTL conditioned on the trait level. The correlation of the effects of these vQTLs between the original and conditional analysis was 0.99 (**Supplementary** Figure 3), consistent with vQTL effects being independent of those for mean trait level.

Across 29 traits, the magnitude of the genetic correlation between trait variance and trait level, as estimated by LDSC^26^, was on average 0.328 (SD=0.24) (**Figure 1c**) and the genetic correlation between trait variance and value was not significant for 21 out of 29 traits after adjusting for multi-testing. Notably, red cell distribution width (rdw) and neutrophil percentage of white cells (neut_p) had significant negative genetic correlations after adjustment for multiple testing, indicating genetic control of trait variance so it is reduced at high levels of rdw or neut_p. Rdw is itself a measure of variation; however, high rdw is an indicator of iron or other nutrient deficiencies, thus our results suggest a potential simultaneous genetic stabilisation when rdw is genetically high. Similarly, high neut_p is an indicator of microbial or inflammatory stress, thus a negative genetic correlation suggests a stabilisation at genetically high neut_p levels.

With many known trait-associated alleles under negative selection^28^, we also assessed the extent to which QTLs for trait variability are under selection. We used Bayes(S)^28^, to compare the selection coefficient (S) between vQTLs and additive QTLs across 29 blood cell traits (**Figure 1d**). We found that, on average S is 1.8 times stronger on trait variance (−0.82, SD=0.07) than trait level (−0.45, SD=0.05) (**Figure 1d**). These results show a much stronger negative selection on blood cell trait variance than on trait level. While it can be difficult to differentiate between negative and stabilising selection, our results are consistent with evolution acting on blood cell traits to remove extreme phenotypes from the population.

We used FUMA^25^ to annotate the lead vQTLs for each trait (**Supplementary Data 1** and **Supplementary Table 6**) and perform a trait enrichment analysis with GWAS Catalog^23^. We found multiple significant overlaps between vQTL and additive QTL related to alcohol consumption. Significant vQTLs (rs191673261 in LD with lead vQTL rs572454376) for platelet crit (pct) were located proximal to *ALDH2*, a well-known gene contributing to alcohol consumption^29^ (**Figure 2a**). We subsequently performed Summary-data-based Mendelian Randomisation (GSMR)^30^ between GWAS of alcohol consumption (as exposure, obtained from Cole et al 2020^31^) and variances of blood cell traits (as outcome). We also used MR-PRESSOR^32^ and MR-weighted median^33^ to validate our results with different assumptions. We did not find statistically significant causal links between alcohol consumption and pct. However, at multi-testing adjusted p < 0.05 level, increased alcohol consumption was genetically predicted to increase variance in mean corpuscular volume (mcv) and mean sphered corpuscular volume (mscv) (**Figure 2b-d**). At nominal significance (p < 0.05 for each of the three MR methods), increased alcohol consumption was genetically predicted to increase variance in red blood cell count (rbc) and neutrophil percentage of white cells (neut_p) (**Figure 2b**). The positive effects of alcohol consumption on neutrophil count (neut) were significant in GSMR (nominal p = 0.014) and MR-PRESSO (nominal p=0.008), but insignificant (nominal p=0.1) in MR-weighted median. Overall, our results support alcohol consumption as affecting particular blood cell trait variances.

**Figure 2.**
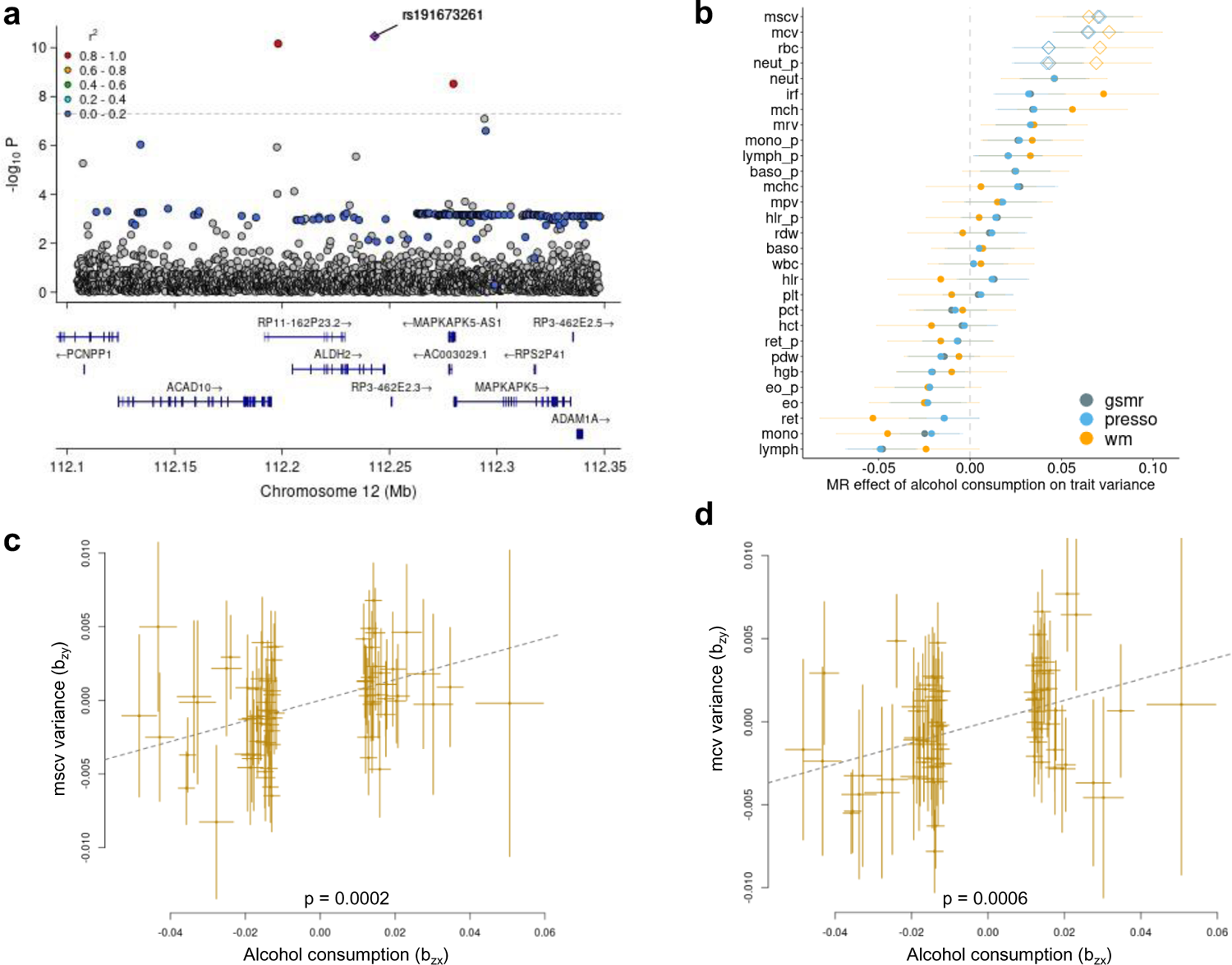
**a**: LocuzZoom plot of variance QTL mapping for platelet crit (pct) variance at ALDH2 gene; **b**: Mendelian randomization (MR) of alcohol consumption on variance of blood cell traits using GSMR^30^, MR-PRESSO (presso)^32^ and weighted-median (wm)^33^. Diamonds: significant in 3 methods; grey dots: p>=0.05; the error bars indicate standard errors; **c**: Effects of MR of alcohol consumption on variance of corpuscular hemoglobin concentration (mscv); **d**: Effects of MR of alcohol consumption on variance of corpuscular volume variance (mcv). Dashed fitted lines indicate the coefficient of Mendelian Randomisation (b_xy_=0.07, se_xy_=0.019 for mscv and b_xy_=0.064, se_xy_=0.0188 for mcv).

FUMA-enabled ANNOVAR^24^ was used to study the enrichment of vQTLs in different functional annotation classes. We found that vQTLs for mean sphered corpuscular volume (mscv), reticulocyte count (ret) and reticulocyte fraction of red cells (ret_p) were significantly enriched in exonic variants related to protein-coding functions (**Supplementary** Figure 4a). However, vQTLs for many other traits were enriched in regulatory regions. For example, vQTLs for mean corpuscular hemoglobin concentration, red blood cell count and hemoglobin concentration (hgb) were enriched for upstream gene regulatory sites. vQTLs for eosinophil count (eo), mean corpuscular hemoglobin and mean corpuscular volume were enriched for downstream regulatory sites of genes. vQTLs for platelet distribution width (pdw) and basophil percentage of white cells (baso_p) were enriched for UTR-3’ sites (**Supplementary** Figure 4a). We used pathway enrichment analyses within FUMA to further investigate whether vQTLs were enriched for gene regulation, finding that vQTLs for mean corpuscular hemoglobin were enriched for many epigenetic regulatory mechanisms including DNA methylation and histone modifications (**Supplementary** Figure 4b).

### Polygenic scores of blood cell trait variance

Polygenic scores are conventionally constructed for differences in trait level. Using the vQTL results from the UK Biobank, we constructed polygenic scores for blood cell trait variance (vPGS) using PRSICE^34^ and the INTERVAL study as an external validation cohort (**Supplementary Table 1**, **Methods**). For conventional PGS we utilised those from Xu et al^7^. Across traits, there was nearly zero Pearson correlation between vPGS and PGS (mean 0.00028, range [-0.018, 0.023]; **Supplementary** Figure 5), consistent with PGS for trait variance being independent from those for mean trait levels.

A potential use of vPGS is to stratify a population by trait variance, thus identifying subgroups where predictive models may have increased performance. For each trait, we stratified individuals into the top and bottom 5% of vPGS. As vPGS were trained to estimate SNP effects on trait variance, individuals with lower or higher vPGS were expected to display less or more variation around the trait mean, respectively. We then compared the correlation of PGSs for each trait between these more (high-vPGS) or less variable (low-vPGS) groups. Across the 27 blood cell traits, we found the less variable group (bottom 5% of vPGS) had a significantly higher PGS-trait correlation than the more variable group (top 5% vPGS) (**Figure 3**). Across all traits, the mean relative difference in PGS-trait correlation (Pearson) between the less and more variable groups was +6.5% [-7%, 18%] (**Figure 3**), with a mean difference of +6.6% [-9%, 19%] for spearman correlation (**Supplementary** Figure 6).

**Figure 3.**
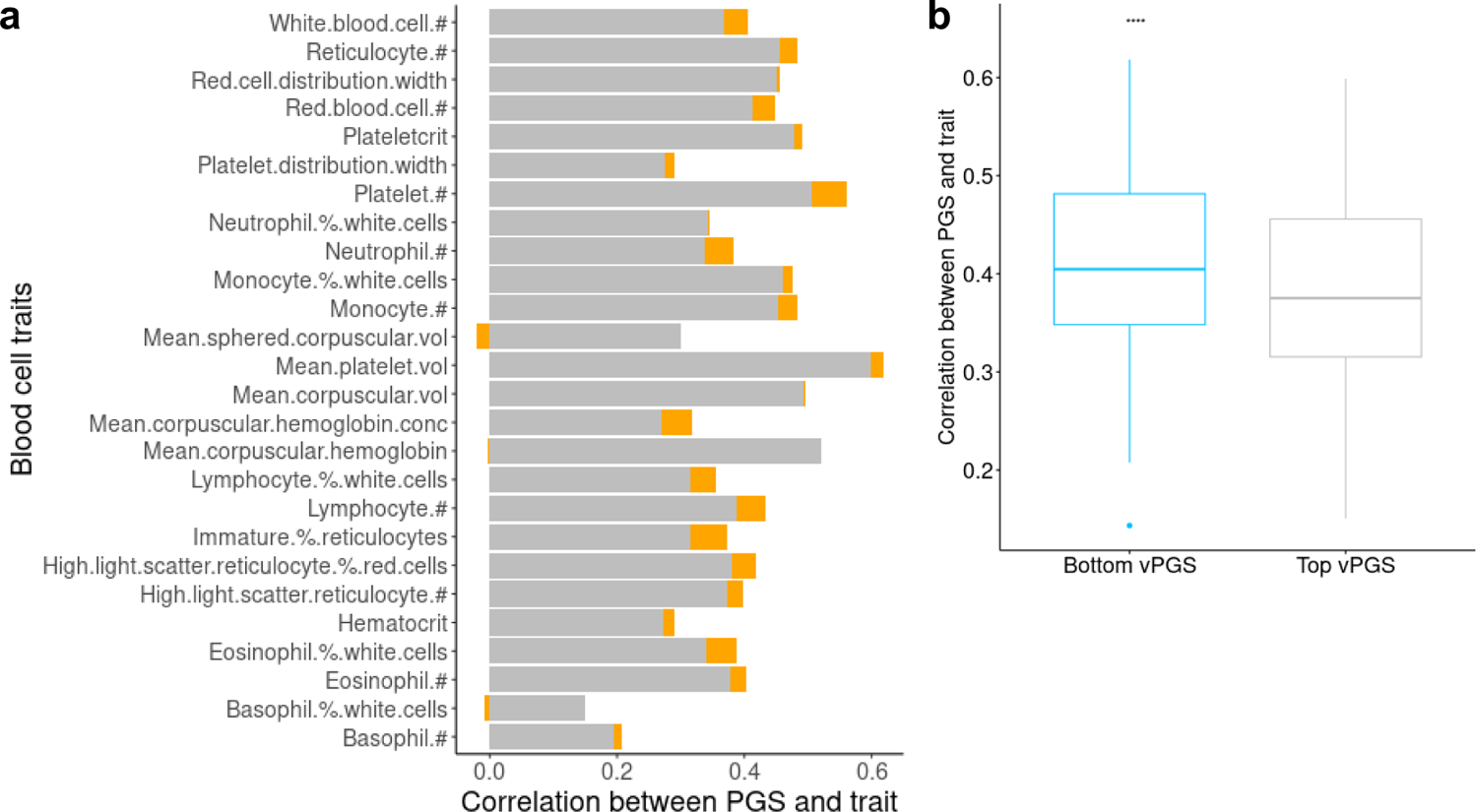
The variation in the accuracy of PGSs for 27 blood cell traits (Pearson correlation) between the top and bottom vPGS groups. **a**: Accuracy of PGS in the top vPGS group (more variable group, grey colour) and the difference (orange) of PGS between the top vPGS group and the bottom vPGS group (less variable group). #: count; % percentage; vol: volume; conc: concentration. **b**: Difference of accuracy of PGS between the bottom and top vPGS groups across 27 blood cell traits. ****: p < 0.0001.

Next, we analysed the effects of interaction between PGS and vPGS for each trait. We found that 6 out of 27 blood cell traits displayed statistically significant (p < 0.05) effects of interaction between PGS and vPGS (**Figure 4a**), suggesting that associations between PGS and blood cell trait level can depend on vPGS (**Figure 4b-c**).

**Figure 4.**
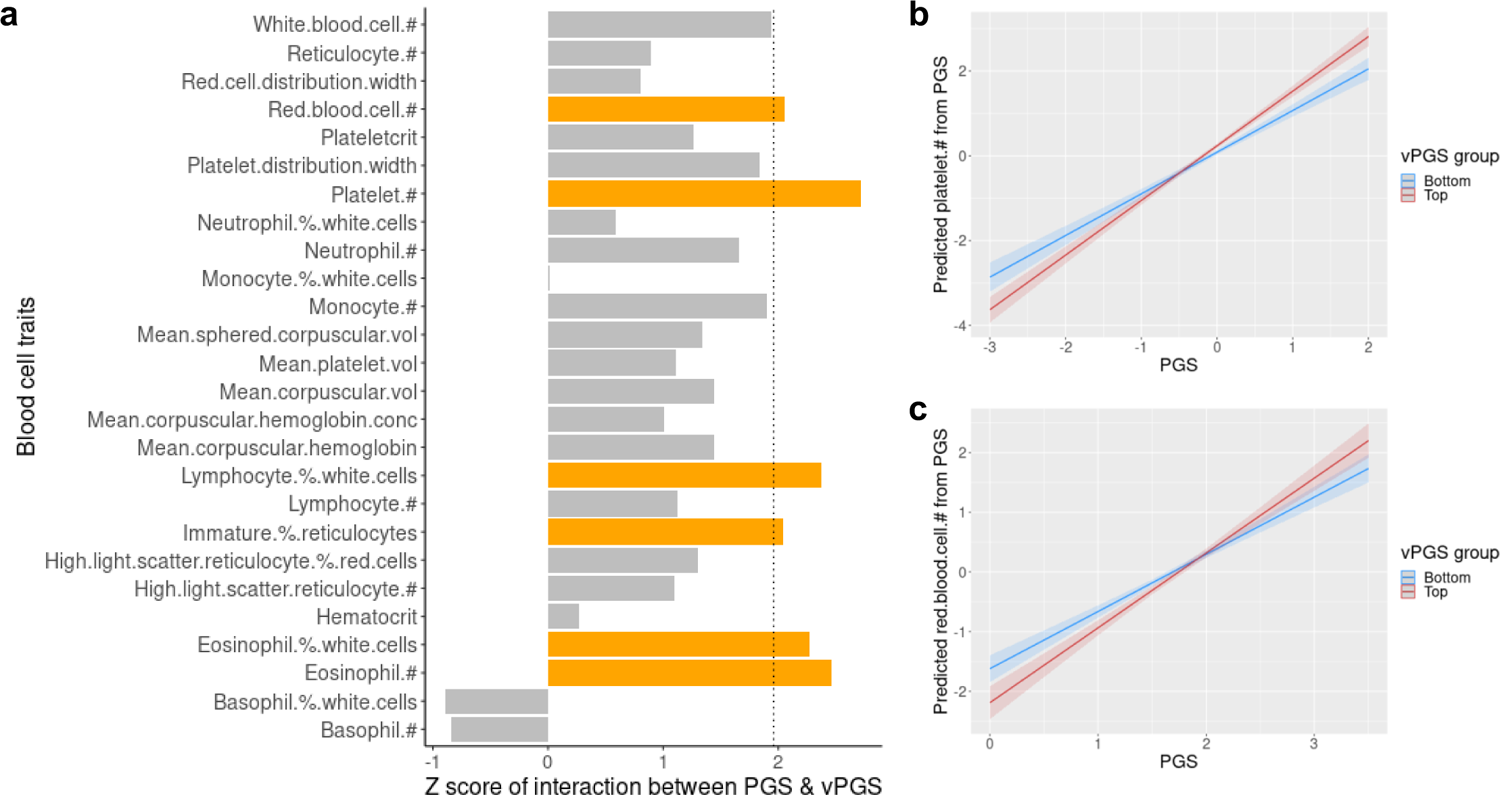
Effects of interaction between PGS and vPGS on blood cell traits. **a**: Effects of interaction across 27 traits in INTERVAL. The vertical dashed line indicates the z-score value = 1.96 which equals p-value = 0.05 and bars with z-score value > 1.96 (p < 0.05) are in orange color. #: count; % percentage; vol: volume; conc: concentration. **b-c**: Examples of visualised effects of interaction for eosinophil percentage of white cells (eo_p) and neutrophil count (neut).

Next, for all INTERVAL individuals, we examined whether adding vPGS to PGS increased the prediction of blood cell trait level. For each blood cell trait, we estimated the difference in the variance explained (*R*^2^) between PGS models with or without vPGS (**Figure 5**, **Methods**). Across all 27 traits, the mean *R*^2^ increase was +1.8% (range [0%, 5%]) and 9 traits showed a statistically significant^35^ increase in *R*^2^ (**Figure 5**, **Methods**). We further tested whether multi-trait vPGSs also increase prediction power^36^, and found that adding multi-trait vPGSs to PGS increased *R*^2^ by a mean of +3.5% (range [0%, 10%]) and the increase was statistically significant in 16 traits (**Figure 5**).

**Figure 5.**
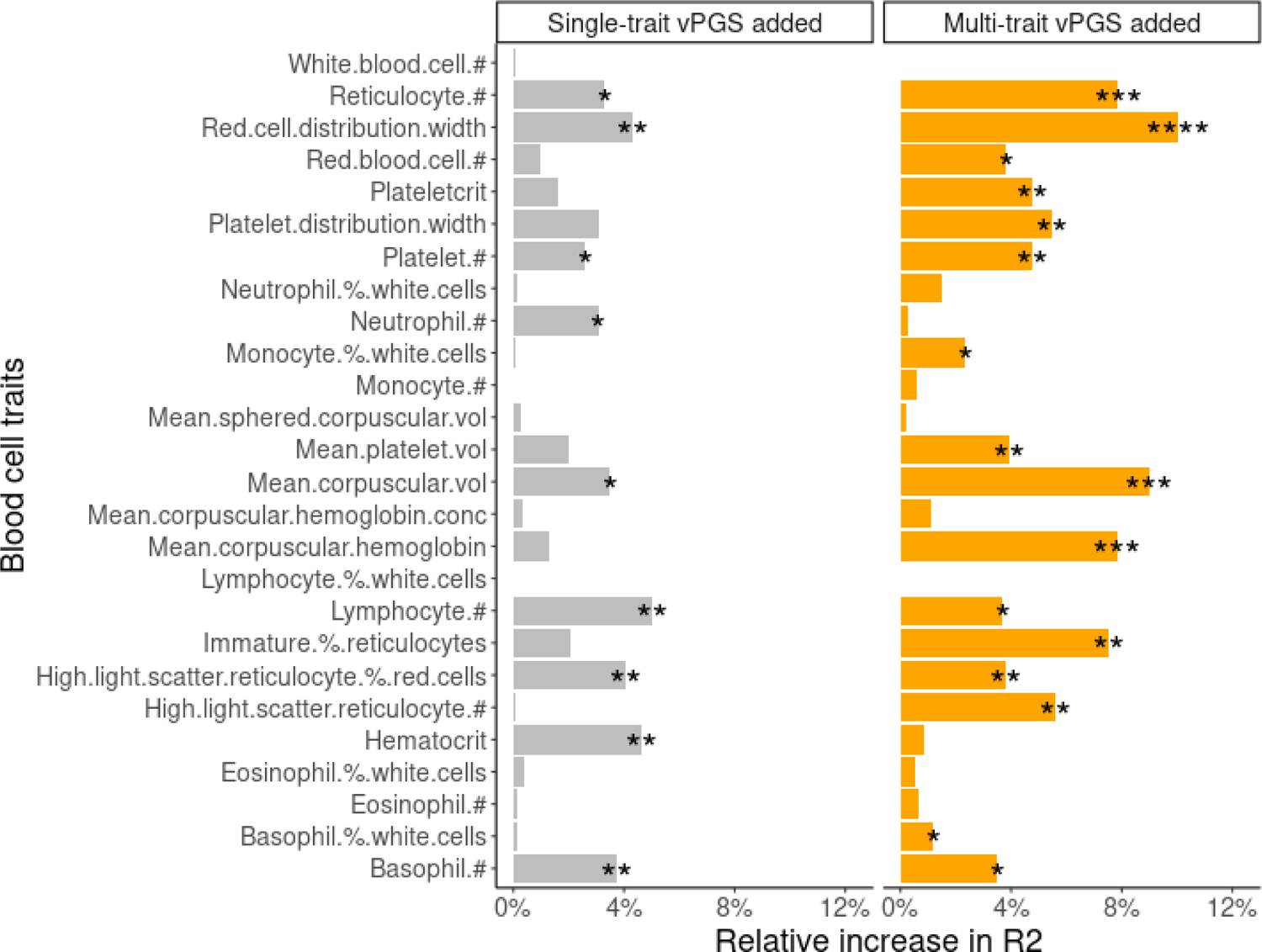
The difference in the variance explained (*R*^2^) between PGS models with or without vPGS. Each bar represents the relative increase in *R*^2^ for the blood cell trait when the PGS model added vPGS. In the left panel, the single-trait vPGS was added to PGS. In the right panel, multi-trait vPGS was added to PGS. #: count; % percentage; vol: volume; conc: concentration. *: p < 0.05; **: p < 0.01; ***: p < 0.001 and ****: p < 0.0001. P-values were estimated by comparing models with and without vPGS using r2redux ^35^.

### Lifestyle effects on blood cell trait variance

To investigate why some individuals have highly variable blood cell trait levels we assessed two major lifestyle factors, namely alcohol consumption and smoking behaviour. We first identified distinct groups of individuals with high or low trait variance in INTERVAL. For the high variability trait group, we identified individuals who were in the top 5% of vPGS for at least 4 blood cell traits and, for the low variability trait group, with individuals in the bottom 5% of vPGS for at least 4 traits (**Methods**, **Figure 6**). Our analysis found that those in the high variability trait group were more likely to be current or previous consumers of alcohol (**Figure 6a**). Further, we applied this analysis to mcv, neut_p and rbc, finding significant causal effects of alcohol consumption in GSMR analyses (**Figure 2a**, mscv not available in INTERVAL). Consistent with the results from GSMR, individuals with high variability in mcv, neut_p and rbc were more likely to be alcohol consumers (**Figure 6b**). These results support the hypothesis that alcohol consumption increases variation in blood cell traits.

**Figure 6.**
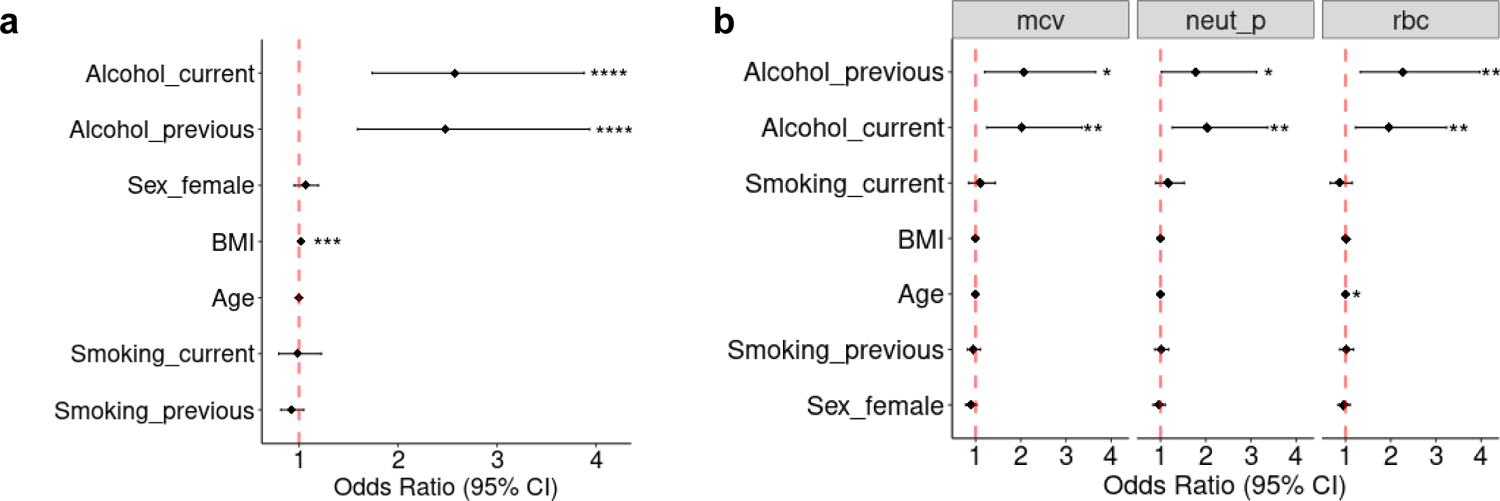
Association between BMI, age, alcohol drinking and smoking and individuals to be genetically variable across blood cell traits in INTERVAL. **a**: an overall estimate across 27 blood cell traits. **b**: Estimates for mean corpuscular volume (mcv), neutrophil percentage of white cells (neut_p) and red blood cell count (rbc) which were significant Mendelian Randomisation analyses. *: p < 0.05; **: p < 0.01; ***: p < 0.001 and **** p < 0.0001.

## Discussion

The analysis of vQTL and vPGS may yield new insights into locus and GxE discovery as well as the use of human genetics for patient stratification. Our study explored vQTL analysis in 29 blood cell traits in the UK Biobank, where the majority (84%) of vQTLs did not overlap with and were largely independent of genetic variants identified in conventional GWAS of trait mean. We investigated the functional annotation, pathway-level associations and selection of vQTLs. The potential utility of using vQTLs to construct vPGS and using the latter to stratify the population into groups of trait variance was demonstrated. Finally, our analysis also showed trait variance to be related to non-genetic factors, finding that alcohol consumption had a putatively causal effect on increasing blood cell trait variances.

Both blood cell trait variance and level display significant negative selection. Stabilising selection of human traits has been reported^14^. However, to our knowledge, negative selection on blood cell trait variance, particularly its strength relative to that on trait level, has not yet been identified. Strong negative selection of blood cell trait variances suggests that extreme blood cell morphologies, which may be indicative of diseases, have not been favoured.

Many vQTLs tagged loci implicated in GxG, GxE and under epigenetic regulation, consistent with previous studies of vQTLs^18,37^. We found blood cell vQTLs tagged genes related to diet. Previous GWAS of diet identified loci related to blood lipids^38^ and glycated hemoglobin^39^ but not to blood cell traits analysed here; however, others have reported that alcohol intake increases mean corpuscular volume independent of the genetic contribution to the level of mean corpuscular volume^40^. In our study, alcohol consumption-related loci significantly overlapped with vQTLs for platelet count, the function of which can be significantly affected by alcohol drinking^41^.

Stratification by vPGS was shown to identify groups with significantly different PGS prediction accuracy, indicating that some groups are intrinsically harder to predict by PGSs than others. Such information may be important for the implementation of PGSs in healthcare. Interestingly, our analysis also found multiple significant interactions between PGS and vPGS, suggesting that the non-additive and GxE components related to PGSs could impact prediction accuracy. These findings are consistent with previous observations^42,43^.

Our results also showed that alcohol consumption and, to some extent increased BMI, were significant contributors to increased genetic variability in blood cell traits. Previously reports have found that blood cell traits can be significantly influenced by alcohol intake^44^ and BMI^45^. However, to our knowledge, this is the first study to report lifestyle risk factors contributing to genetically predicted variation in blood cell traits.

In conclusion, our study provides an in-depth analysis of human genetic effects on the variance of blood cell traits, including the discovery of loci and strong negative selection, improved genomic prediction and stratification, and identification of GxE. vPGSs may provide a generalisable approach to incorporate individual differences to improve trait and disease risk prediction. This study demonstrates that there is substantive human biology and potential clinical utility in studying trait variances alongside conventional studies of trait means.

## Methods and Materials

### Study Cohorts and Methods

#### UK Biobank

The UK Biobank^46,47^ (https://www.ukbiobank.ac.uk/) is a cohort including 500,000 individuals living in the UK who were recruited between 2006 and 2010, aged between 40 and 69 years at recruitment. Ethics approval was obtained from the North West Multi-Center Research Ethics Committee. The current analysis was approved under UK Biobank Project 30418. The participants with the measurements of the 29 blood cell traits and who were identified as European ancestry based on their genetic component analysis were included in our study. The detailed sample sizes used for vQTL detection were shown in **Supplementary Table 1**.

#### INTERVAL Study

INTERVAL^23^ (https://www.intervalstudy.org.uk/) is a randomised trial of 50,000 healthy blood donors, aged 18 years or older at recruitment. The participants with measurements of the 27 considered blood cell traits were included in our study. The detailed sample sizes were shown in **Supplementary Table 1**. All participants have given informed consent and this study was approved by the National Research Ethics Service (11/EE/0538).

#### Data quality control

For trait levels of 29 blood cell traits in the UK Biobank and matching 27 traits in the INTERVAL, we adopted previously established protocols for quality controls^5-7^ to adjust technical and other confounders and the first 10 genetic principal components. For trait levels, adjusted technical variables include the time between venepuncture and full blood cell analysis, seasonal effects, center of sample collection, the time-dependent drift of equipment, and systematic differences in equipment; other adjusted variables included sex, age, diet, smoking and alcohol consumption. Quality control and imputation of the genotype data have been described previously^5,47^, which filtered the samples to the European ancestry only.

#### vQTL analysis

Genome-wide analysis of vQTL used Levene’s test. As detailed in^11,15^, the test statistic of Levene’s test is:

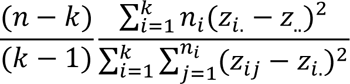

where *n* is the total sample size, *k* is the number of groups (k = 3 in vQTL analysis), *n*_*i*_ is the sample size of the *i*th group (one of three genotypes), *Z*_*ij*_ is the absolute difference between the phenotype value in sample *j* from genotype and the median value in genotype *i*, *Z*_*i*._ is the average *Z* value in genotype *i*, and *Z*_.._ is the average *Z* value across all samples. OSCA-implemented Levene’s test also provides beta and se estimates based on p-value and minor allele frequency^15^ and the beta estimates were used to construct vPGSs described later.

We estimated the study-wise significance for vQTL as 4.6 × 10^-9^ = 5 × 10^-8^ / 10.2 where 10.2 is the effective number of traits analysed in the study. The effective number of traits is estimated using 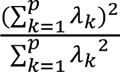, where λ_1_.. λ_*p*_ is principal component variances or the ordered eigenvalues^15,17^. To identify lead vQTL with relative independence, we used plink-clumping^48^ using a p-value threshold of 4.6 × 10^-9^ *r^2^* <0.01 and window size of 5000kb (the same parameter used by^15^). The LD analysis between vQTL and lead QTL reported by Vuckovic et al ^6^ used plink 1.9 with the function of --ld. The novel vQTL was defined as those lead vQTL after clumping with GWAS p-value > 4.6 × 10^-9^ in Vuckovic et al and with LD-r^2^ < 0.8 with lead QTL reported by Vuckovic et al. For vQTL mapping results of each trait, we used LDSC^26^ to estimate lambda-GC and intercept to check inflation. We also used FUMA^49^ to annotate significant vQTL for each trait with default settings. Results from FUMA functions of SNP2GENE and GENE2FUNC were presented in the results.

To explore the potential causal relationships between alcohol consumption and blood cell trait variances, we used GSMR^30^ to discover the causal relationships and used MR-PRESSO^32^ and weighted-mean implemented in MendelianRandomisation^33^ as validation. The GWAS summary data for alcohol consumption was obtained from Cole et al^31^. Default settings for nominated software were used and SNPs with p-value < 5e-8 and r^2^ < 0.05 were used in the analysis. Significant results were defined as the multi-testing adjusted p-value from GSMR < 0.05 and the nominal significance was defined as the Mendelian Randomisation had raw p-value < 0.05 in all 3 methods.

#### Analysis of vPGS and PGS

PGS trained using the elastic net from Xu 2022 et al^7^ was used. For training vPGS, we followed the protocol described by Miao et al^16^ reported successful implementation of vPGS for BMI using PRSICE^34^, we used the same procedure described by Miao et al to construct vPGS in the INTERVAL using PRSICE, i.e., –clump-p1 1 –clump-p2 1 –clump-r^2^ 0.1 and –clump-kb 1000. When vPGS was computed for each trait, they were used to rank INTERVAL individuals where the top and bottom 5% of individuals were stratified. As vPGS was trained based on SNP effects on phenotypic variance, i.e., the extent to which the individual measurement deviates from the mean, vPGS was expected to genetically predict such variation of individuals for the corresponding trait. Therefore, individuals ranked in the top 5% of vPGS were called the genetically more variable group and individuals ranked in the bottom 5% of vPGS were called the genetically less variable group. Then, for each trait, within the more variable and less variable groups, we estimated the PGS accuracy, i.e., the correlation between PGS and the corresponding trait. We then compared the PGS accuracy between the more variable and less variable groups for each trait and the relative increase was calculated as 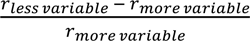 where *r_less variable_* is the PGS accuracy in the less variable group defined by vPGS and *r*_*more variable*_ is the PGS accuracy in the more variable group defined by vPGS.

The effects of interaction between PGS and vPGS on the corresponding trait in INTERVAL were tested on corrected blood cell traits (described above). As the traits were already corrected for covariates, only the main effects and interaction of PGS and vPGS were fitted for each blood cell trait in the lm() function in R: *y* = *PGS* + *vPGS* + *PGS* ∗ *vPGS*, where y was each of the blood cell trait. The effects of interaction on specific traits (e.g., eo_p and neut) were visualised using the function of plot_model in the R package sjPlot (version 2.8.15).

To evaluate if adding vPGS improves PGS model predictability, we tested two sets of vPGS, where one set is the original single-trait vPGSs for 27 traits computed by PRSICE, and the other set is estimated using the multi-trait BLUP (SMTpred^36^) combining information from single-trait vPGSs. Following the instructions from https://github.com/uqrmaie1/smtpred, we used the LDSC^26^ wrapper (ldsc_wrapper.py) with default options in SMRpred to estimate the genetic parameters for each trait which are required inputs by the multi-trait BLUP. Then, the script smtpred.py was used by default options with the estimated genetic parameters to combine single-trait vPGSs to construct multi-trait vPGSs. Then, we used r2redux^35^ to quantify the difference in variance explained (*R*^2^) between PGS models with and without vPGS. As described by Momin et al^35^, r2redux can powerfully detect *R*^2^ differences between models for the out-of-sample genomic prediction which is suitable to our case where the PGS and vPGS models were trained in the UK Biobank and predicted into INTERVAL. We followed the instructions provided by (https://github.com/mommy003/r2redux) to compare the *R*^2^ of models with vPGS and without PGS using the nested method and obtained p-values testing the significance of the increase in *R*^2^ when adding vPGS. The relative increase in *R*^2^ was expressed as the absolute difference in *R*^2^ divided by the heritability estimated using LDSC^26^.

To characterise the individuals that were identified as genetically variable across traits, we first counted the number of times (out of 27 blood cell traits) an individual was ranked in the top 5% by PGS for each trait. We also counted the number of times an individual was ranked in the bottom 5% by PGS for each trait. We then identified 2,465 individuals who always ranked in the top 5% vPGS, and 2,362 individuals who always ranked in the bottom 5% vPGS across multiple blood traits. Individuals in the top group were ranked in the top 5% vPGS for 4 to 17 traits with a mean of 5 and individuals in the bottom group were ranked in the bottom 5% vPGS for 4 to 23 traits with a mean of 9. Then, the top group was labelled as 1 and the bottom group was labelled as 0 and this 0/1 vector was analysed as a binary outcome for a logistic regression analysis against lifestyle factors: *y* = *age* + *sex* + *BMI* + *smoking_status* + *drinking_status*, where the average age is 46.1 (SD=14.3) and the average BMI is 26.2 (SD=4.6); for sex, there are 2,419 women; for smoking status, there are 2,728 people never smoked, 378 current smokers, 1,634 previous smokers and 87 with no answers; for alcohol drinking status, there are 118 who never drunk, 4,178 current drinkers, 323 previous drinkers and 208 with no answers. The logistic regression used the function glm() in R and for sex the male was set to the reference level, for smoking the level of never smoked was set to the reference and for drinking the level of never drunk was set to the reference. The same analysis was also applied to individual blood cell traits of mean corpuscular volume (mcv), neutrophil percentage of white cells (neut_p) and red blood cell count (rbc) which were significant in Mendelian Randomisation analyses.

## Data availability

Full summary statistics of vQTL mapping are available via the GWAS Catalog (https://www.ebi.ac.uk/gwas/) under the accession number NNNNNNNNN (*to be generated upon acceptance of peer-reviewed manuscript*). All data described are available through the UK Biobank subject to approval from the UK Biobank access committee. See https://www.ukbiobank.ac.uk/enable-your-research/apply-for-access for further details. INTERVAL study data from this paper are available to bona fide researchers from and information, including the data access policy, is available at http://www.donorhealth-btru.nihr.ac.uk/project/bioresource.

## Code availability

The manuscript does not produce original code. vQTL mapping used OSCA: https://yanglab.westlake.edu.cn/software/osca/#Overview; genetic correlation analysis used LDSC: https://github.com/bulik/ldsc; pleiotropy analysis: https://github.com/rondolab/HOPS. Mendelian randomisation used GSMR: https://yanglab.westlake.edu.cn/software/gsmr/, MR-PRESSO: https://github.com/rondolab/MR-PRESSO and MendelianRandomisation: https://cran.r-project.org/web/packages/MendelianRandomization/index.html; Analysis of selection used GCTB-BayesS: https://cnsgenomics.com/software/gctb/#SummaryBayesianAlphabet; vPGS analysis used PRSICE: https://choishingwan.github.io/PRSice/ and plink2: https://www.cog-genomics.org/plink/2.0/; multi-trait GBLUP used SMTpred: https://github.com/uqrmaie1/smtpred; significance tests of *R*^2^ increase used r2redux: https://github.com/mommy003/r2redux; logistic regression analysis used glm(): https://www.rdocumentation.org/packages/stats/versions/3.6.2/topics/glm.

## Competing interests

M.I. is a trustee of the Public Health Genomics (PHG) Foundation, a member of the Scientific Advisory Board of Open Targets, and has research collaborations with AstraZeneca, Nightingale Health and Pfizer which are unrelated to this study.

## Funding

The authors are grateful to UK Biobank for access to data to undertake this study (Projects #30418). This work was supported by core funding from the: British Heart Foundation (RG/18/13/33946) and NIHR Cambridge Biomedical Research Centre (NIHR203312), Cambridge BHF Centre of Research Excellence (RE/18/1/34212) and BHF Chair Award (CH/12/2/29428) and by Health Data Research UK, which is funded by the UK Medical Research Council, Engineering and Physical Sciences Research Council, Economic and Social Research Council, Department of Health and Social Care (England), Chief Scientist Office of the Scottish Government Health and Social Care Directorates, Health and Social Care Research and Development Division (Welsh Government), Public Health Agency (Northern Ireland), British Heart Foundation and Wellcome. X.J. was funded by the British Heart Foundation (CH/12/2/29428) and Wellcome Trust (227566/Z/23/Z). S.C.R. was funded by a British Heart Foundation (BHF) Programme Grant (RG/18/13/33946). S.C.R. was also funded by the National Institute for Health and Care Research (NIHR) Cambridge BRC (BRC-1215-20014; NIHR203312) [*]. Y.X. and M.I. were supported by the UK Economic and Social Research Council (ES/T013192/1). M.I. and S.L. were supported by NIH U24 (5U24HG012542-02). M.I. is supported by the Munz Chair of Cardiovascular Prediction and Prevention and the NIHR Cambridge Biomedical Research Centre (NIHR203312) [*]. *The views expressed are those of the authors and not necessarily those of the NIHR or the Department of Health and Social Care.

## Author contributions

R.X. and M.I. conceived of the study. R.X. performed the analyses with assistance from Y.X. and M.I.. R.X., M.I., F.T., Y.L., C.B.E., X.J., S.R., S.A.L., and Y.X. drafted and revised the manuscript. All authors read and approved the final version of the manuscript.

## Acknowledgements

We thank Dr. Emmanuela Bonglack and Dr. Xilin Jiang, for their insightful comments. The authors are grateful to Prof. Michael Goddard for discussions on the selection of vQTL.

## Notes

### Author Declarations

All data described are available through the UK Biobank subject to approval from the UK Biobank access committee. See https://www.ukbiobank.ac.uk/enable-your-research/apply-for-access for further details. INTERVAL study data from this paper are available to bona fide researchers from and information, including the data access policy, is available at http://www.donorhealth-btru.nihr.ac.uk/project/bioresource.

