## Supplementary Figures 1-6 for "Genome-wide analyses of variance in blood cell phenotypes provide new insights into complex trait biology and prediction"

This file contains:
Supplementary Figures 1-6

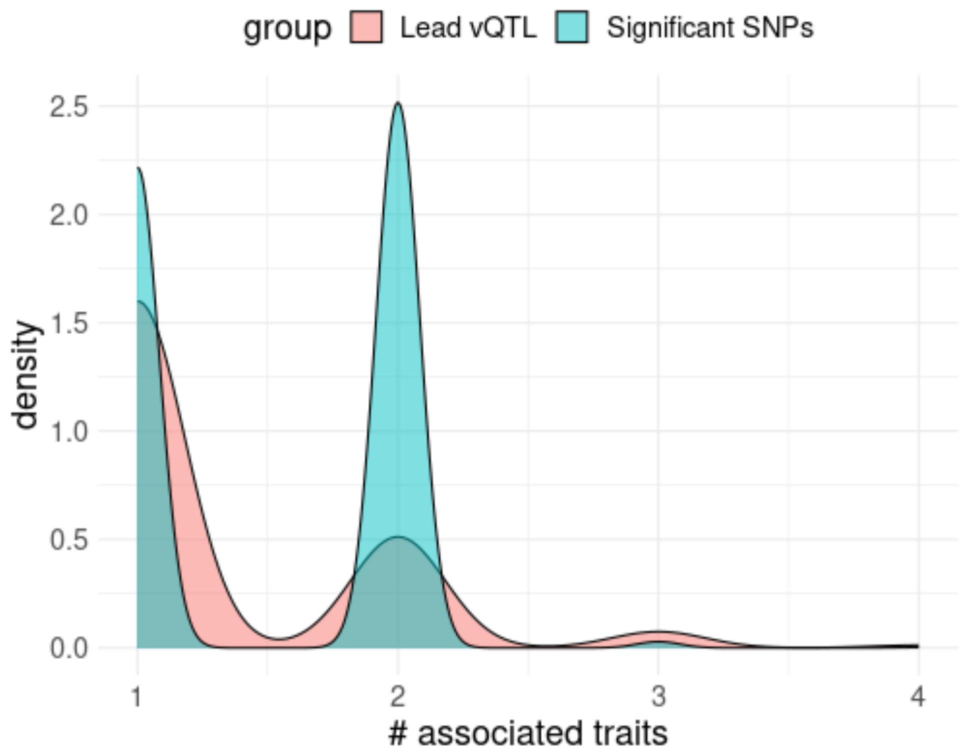


**Supplementary Figure 1**. Pleiotropy of vQTL. Lead QTLs were clumped with LD-$r^2 < 0.01$. Significant SNPs (not clumped) were associated with the variance of any traits with $p < 4.6 \times 10^{-9}$.

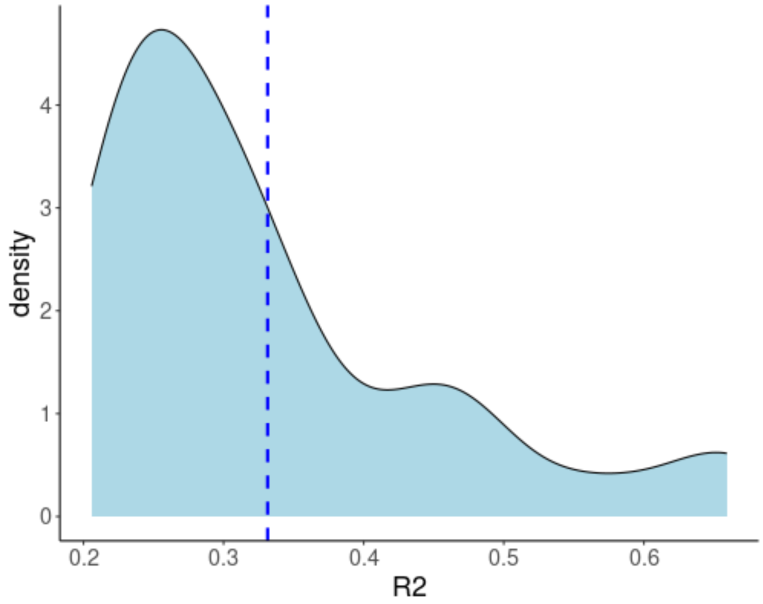


**Supplementary Figure 2**. Distribution of LD-r2 between novel lead vQTL identified in the current study and lead SNPs reported by Vuckovic et al 2020[1].

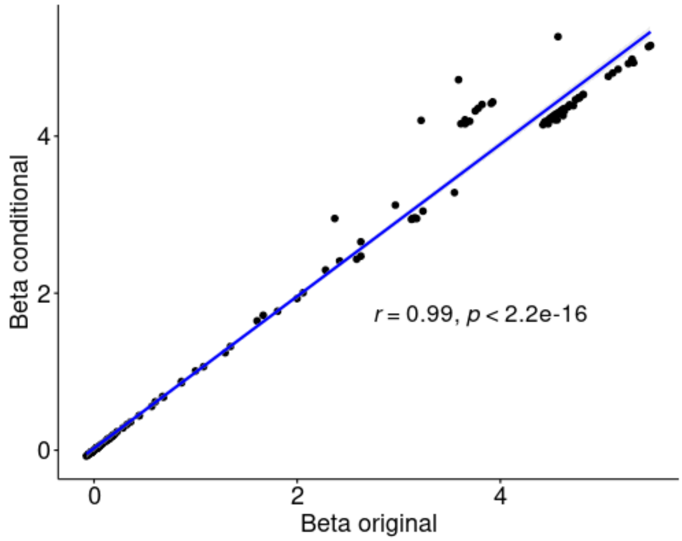


**Supplementary Figure 3**. Correlation of effects of 176 vQTL between original analyses and conditional analysis on the trait level.

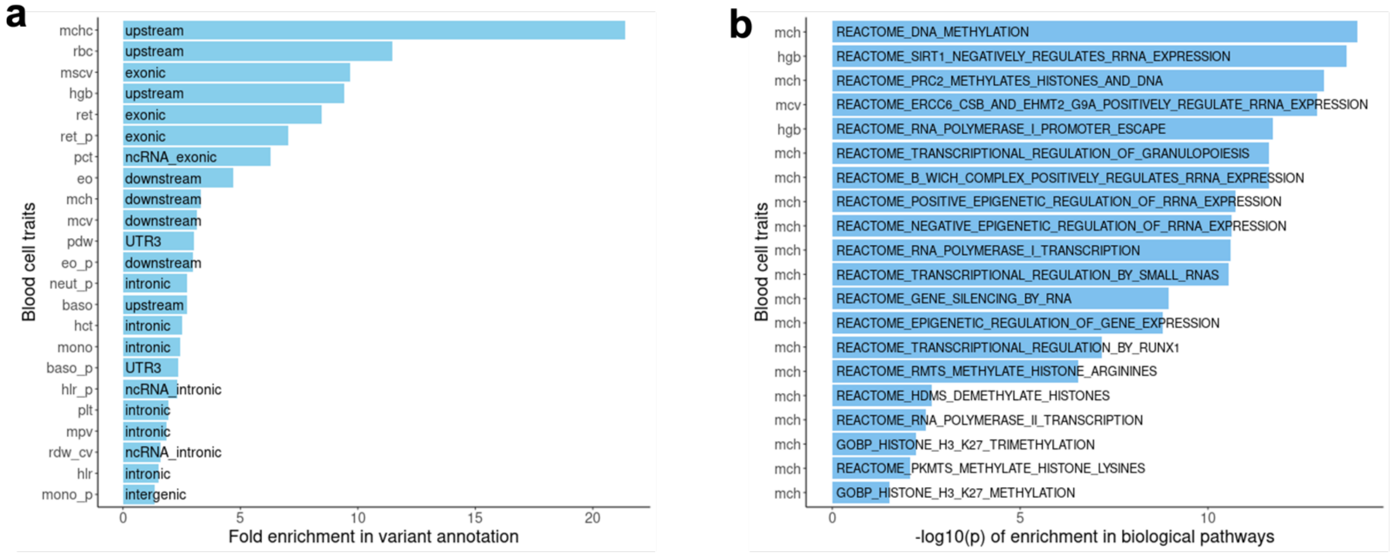


**Supplementary Figure 4. a**: Best enrichment of class of functional consequences (ANNOVAR) in lead vQTLs for each blood cell trait. **b**: Best enrichment of biological pathways related to gene regulation in lead vQTLs for each blood cell trait. All enrichment analyses were conducted using FUMA and only enrichments with multi-testing adjusted p-value < 0.05 are shown.

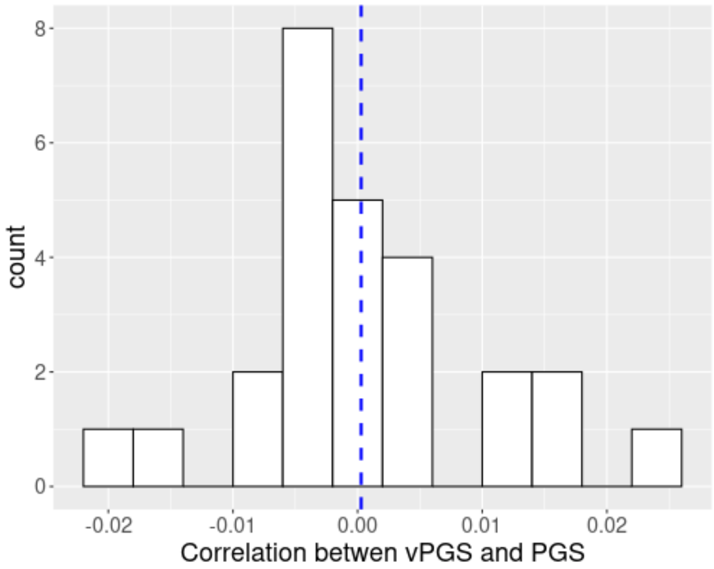


**Supplementary Figure 5**. Distribution of correlation between vPGS and PGS across 27 traits in INTERVAL. The blue line indicates the mean ($r$ = 0.00028 ) of correlation across traits.

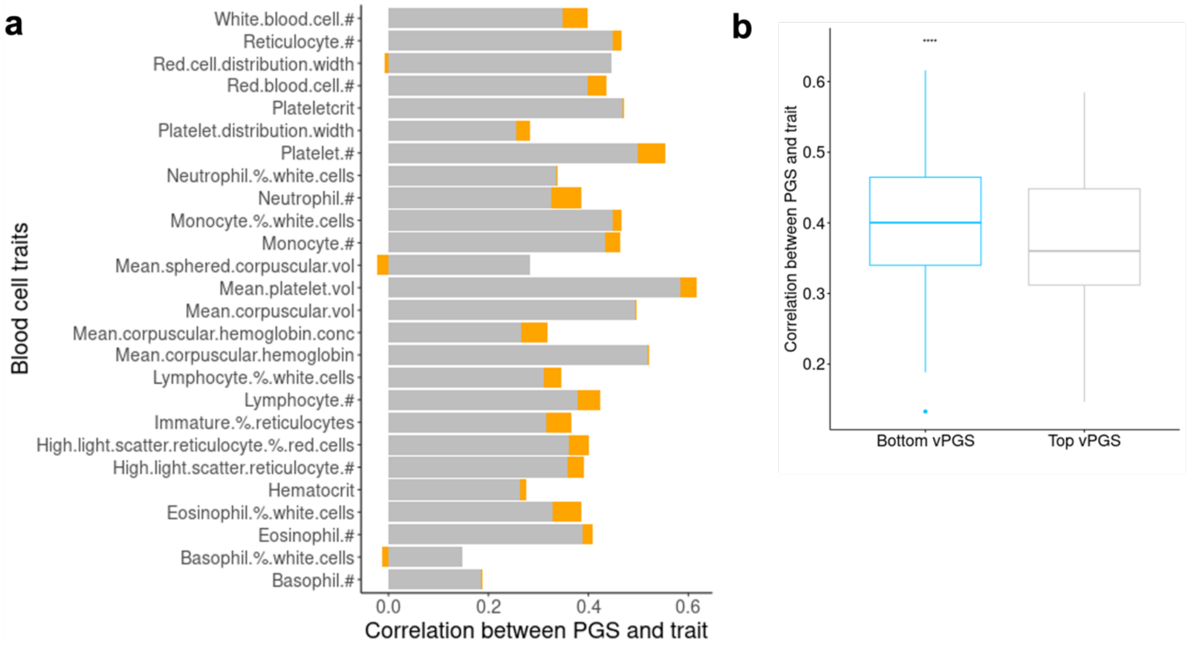


**Supplementary Figure 6**. The variation in the accuracy of PGSs for 27 blood cell traits (spearman correlation) between the top and bottom vPGS groups. **a**: Accuracy of PGS in the top vPGS group (more variable group, grey colour) and the difference (orange) of PGS between the top vPGS group and the bottom vPGS group (less variable group). #: count; % percentage; vol: volume; conc: concentration. **b**: difference of accuracy of PGS between the bottom and top vPGS groups across 27 blood cell traits. ****: $p < 0.0001$.
